# The Impact of Clinical Audits on Improving the Effectiveness of Type 2 Diabetes Mellitus (T2DM) CARE in Primary Health Centers. A Comprehensive Pre-post analysis through Multi-layered Intervention: The ICAE-DM CARE study protocol

**DOI:** 10.1101/2024.10.30.24316448

**Authors:** Ansif Pallath Majeed, Niyaz Panakaje, Shajitha Thekke Veettil, Kiran Harikumar, Hanan Al Mujalli, Abdul Ali Shah, Noora Jassim AlKubaisi, Ahmed Sameer Alnuaimi

## Abstract

**Background:** Conducting clinical reviews is integral to the continuous process of enhancing healthcare quality by identifying specific areas or aspects within medical services and clinical practices. These reviews involve measuring a clinical outcome or process against established evidence-based standards, aiming to spotlight discrepancies between current practice and these benchmarks to facilitate improvements in care quality. Notably, clinical reviews rely on the exceptional skill of the driving force, underscored by key elements: the clinical expertise of participants, result confidentiality, and an objective strongly linked to the professional ‘quality’.

The worldwide prevalence of type 2 diabetes mellitus (T2DM) is on the rise, especially in economically developing nations. Current research conducted in Qatar indicates a significant prevalence of diabetes mellitus within the adult population, alongside a considerable proportion of pre-diabetes cases that are likely to elevate the incidence of diabetes mellitus in the coming years. This information underscores the critical need for effective diabetes management at the primary care level, particularly for individuals with pre-diabetes. A comprehensive systematic assessment and intervention are essential to enhance diabetes care.

**Methods:** The proposed research will adopt a quasi-experimental design, which includes a baseline cross-sectional situational analysis conducted across 31 Primary Health Care Centers (PHCCs) prior to the intervention. The study will focus on patients aged 18 and older who have been diagnosed with Type 2 Diabetes Mellitus (T2DM), encompassing both newly diagnosed individuals and those in follow-up care, while excluding pregnant patients. A random sampling method will be utilized to ensure a representative sample size of 450 patients from the last three months of diabetes consultations. Following the identification of gaps in the situational analysis, an intervention will be implemented, after which a post-intervention cross-sectional study will be carried out using the same sample as the baseline to evaluate changes in the measured parameters. Additionally, a cohort study will be performed through a telephonic survey of a random sample of 60 patients, both before and after the intervention, to assess changes from the patients’ perspective.

**Discussion:** A comprehensive assessment and intervention are essential to evaluate the quality of diabetes care delivered at primary health centers. The ICAE-DM CARE study aims to provide an effective situational analysis, which will include the identification of gaps and the underlying causes of these gaps in the DM care using suitable quality improvement tools. The study will also showcase appropriate intervention strategies to improve practice. Furthermore, the implications of this process will be thoroughly examined. This study design will closely reflect a practical understanding of healthcare quality management in diabetes care, which can be applied in similar settings.

## Background

Clinical Audit as a process for health care quality improvement and patient’s safety has been fully established in hospitals environment. It is a quality improvement process that seeks to improve patient care and outcomes through systematic review of care against the explicit criteria and implementation of change [1]. Conducting a clinical audit has many benefits including improving patient care, demonstrating the benefits of best practice to others, making more effective use of time, increasing number of satisfied patients, helping to advance the practice, identifying areas for making practice more efficient and providing useful evidence of continuing professional development activity and evidence-based practice [2].

Each clinical audit has potential to bring different results, sometimes, gratification of being compliant to the best available practice or an eye-opener on the need of an improvement in the practice. When gaps in the practice have been noticed then an audit becomes indispensable to identify the compliance and take actions to reduce or bridge the gaps [1]. This shows the transparency and Clinical auditing as a quality improvement tool will improve overall patient care [2].

The execution of the clinical audit process in type 2 diabetes management has shown positive results. The study revealed that the interventional audit helped to achieve positive results in managing not only glycaemia but also blood pressure [3]. It has also improved patient satisfaction with the care provided [3]. The execution of clinical audits to improve adherence to diabetes care also worked in a primary care setting. With the help of the audit cycle, a significant improvement was shown in the adherence to the guidelines [4]. Certain studies also used clinical audit as a quality improvement tool to measure the appropriateness of diabetes screening and care provided [5].

Application of the clinical audit process for the management of chronic disease was an eye-opener for certain hospitals to understand their deficiencies. A study conducted in a Thai secondary hospital revealed their hypertension management needed to be strengthened. There is a need for adherence to the guidelines, further training for the staff, and a need to simplify the treatment protocols discovered through this quality improvement process [6]. Suggestion for specialized chronic disease clinic and longer follow up visits to achieve sustained improvement was another upshot of a clinical audit [7].

There are not many studies demonstrating the impact of a complete audit cycle, especially on chronic disease care. This study aims to evaluate and enhance the management of diabetes, including related clinical documentation. The influence of clinical audits on improving chronic disease care will be evaluated with the help of pre-intervention and post-intervention. In the pre-intervention, current practices will be evaluated and reported. And, in the post-intervention, changes in care, if any, will be measured, and the impact of the intervention will be assessed. This study aims to answer whether a clinical audit can improve diabetes care and related documentation in Primary Care Settings. This paper will outline the entire process trajectory as defined by the ICAE-DM CARE study protocol.

## Study Rationale

Conducting clinical reviews is integral to the continuous process of enhancing healthcare quality by identifying specific areas or aspects within medical services and clinical practices [8]. These reviews involve measuring a clinical outcome or process against established evidence-based standards, aiming to spotlight discrepancies between current practice and these benchmarks to facilitate improvements in care quality [8]. Notably, clinical reviews rely on the exceptional skill of the driving force, underscored by key elements: the clinical expertise of participants, result confidentiality, and an objective strongly linked to the professional ‘quality’ [8].

From a strategic standpoint, a clinical review follows a ‘quality circle’ approach: selecting a topic, establishing shared and measurable criteria and standards, evaluating current clinical practices— particularly in terms of processes or outcomes—suggesting and implementing improvements, and subsequently restarting the cycle. This process involves comparing current practices against well-defined standards [9], always with the ultimate goal of enhancing patient care. Achieving this goal entails various actions: (1) fostering a culture of clinician engagement, (2) problem-solving, (3) standardizing professional conduct, and (4) bridging the gap between theoretical standards and real-life application.

Effective clinical audits hinge on implementing recommendations derived from the audits within care settings. Research indicates that organizational barriers to audit implementation stem from the lack of collaboration between clinicians and managers [10]. Disparities in perspectives between clinicians and management, along with ambiguous lines of authority and accountability among clinicians, often create confusion and inertia regarding responsibility for implementing changes. Negotiating changes within the wider hospital exacerbates these challenges [10].

While the Primary Health Care Corporation (PHCC) in Qatar has integrated the clinical audit process as a clinical effectiveness tool since 2012, conducting an annual average of 20 to 25 clinical audits, its specific effectiveness in chronic disease care remains unexplored. Through two comprehensive studies encompassing pre- and post-intervention assessments, our aim is to ascertain the effectiveness of clinical audits in improving diabetes care and associated documentation which is in accordance with PHCC guideline.

## Methods

### Aim

The ICAE-DM CARE study aims to determine the effectiveness of clinical audits in enhancing. T2DM care, related documentation, and patient satisfaction at the primary care level.

### Study settings

Qatar is an Arab nation located on a peninsula. In recent years, the country has made substantial investments in establishing a universally accessible publicly funded healthcare system for both Qatari citizens and non-Qatari residents. The Primary Health Care Corporation (PHCC) serves as the largest provider of primary care in Qatar, offering comprehensive, integrated, and coordinated health services that prioritize individual needs within the community, with an emphasis on disease prevention, promoting healthy lifestyles, and enhancing overall wellness. This research will utilize the data stored by Primary Health Care Centers (PHCCs) as well as information from service users. The review of guidelines will be conducted via online access to the PHCC intranet. Additionally, a review of medical records will be performed using the clinical information system (CIS) of the PHCCs, and patient satisfaction will be assessed through telephone interviews.

### Study design and population

The proposed study will employ a quasi-experimental design for the intervention component and a cohort design for the patient survey, utilizing a random sample of patient records from all health centers within the PHCC. The focus will be on individuals aged 18 and above who have received a diagnosis of Type 2 Diabetes Mellitus (T2DM). Exclusions from the study will include pregnant individuals, telephonic consultations, and medication refill encounters. Additionally, for the telephonic interviews, individuals will be excluded if they are unable to communicate via phone or provide verbal consent.

### Sample Selection

A simple randomization technique will be employed to obtain a representative sample size suitable for a clinical audit concerning diabetes encounters over the past three months at 31 health centers. Taking into account a 5% margin of error, a 95% confidence interval, and a 50% response distribution, the sample size for the cross-sectional study has been established at 450 to account for potential non-responses. For the cohort phase through a telephonic patient survey will be conducted during the pre- and post-intervention phases. A randomly selected sub-sample will be taken to do this survey. A randomly selected 60 cases will be undergo telephonic survey. Same sample size will be taking for pre, and post intervention phase the study. In the cohort phase, a telephonic patient survey will be administered in both the pre- and post-intervention phases. A randomly selected sub-sample will be utilized for this survey, comprising 60 cases that will undergo the telephonic survey. An equivalent sample size will be maintained for both the pre-intervention and post-intervention stages of the study.

### Participant recruitment

For the first cross-sectional study, a random sample of 450 cases will be drawn, in addition to 60 cases for the cohort, from the electronic medical record system of PHCCs, utilizing their specific health record numbers. In the subsequent cross-sectional study, post-intervention, a target sample of 450 cases will again be randomly selected, along with 60 cases for the second cohort, to assess the changes in practice.

### Estimated study timeline

We intend to initiate data collection for Phase 1 (the pre-intervention phase) in early December 2024, which will involve medical record review and conducting telephonic interviews with patients. We anticipate that this phase will be completed within a three-month timeframe. This will be succeeded by a gap analysis and the execution of an action plan (Phase 2), which is projected to take approximately ten months. Finally, a post-intervention analysis (Phase 3) will be conducted. We estimate that the entire study will be concluded by the end of 2025.

### Data collection

Experienced clinical auditors will do the medical record review and data will record in a customize created excel sheet which having all the audit criteria derived from the T2D guideline. The variables will be the audit criteria derived from the corresponding guideline.

Risk Assessment: Blood pressure, weight/BMI, lipids, smoking status, and serum creatinine.

- HbA1c test reviewed, and place ordered for blood workup (if recommended).
- HbA1c results
- Continued or modified treatment based on the HbA1c results.
- Medication refill orders
- Preventive health education is offered to diabetes patients on lifestyle changes.
- Follow-up visits scheduled.
- Vaccination, foot examinations, and annual eye screenings should be provided.
- Appropriate referral to secondary care.

A review of the current practices and their compliance with the PHCC Clinical Practice Guidelines for the Diagnosis and Management of Type 2 Diabetes in Adults will be performed, with the results compiled in an Excel sheet. Information will be sourced from the CIS system by navigating through the Power Chart and additional sections.

A patient satisfaction survey will be executed via telephone, utilizing a questionnaire specifically designed for this purpose and data will record in a customize created excel sheet which having all the measuring parameters. The creation of the questionnaire will be based on the key performance indicators derived from the clinical practice guidelines. This approach has been routinely integrated into clinical auditing, employing questions that have been developed and validated by the Department of Clinical Effectiveness.

Following the identification of gaps in the first cross-sectional study, a comprehensive action plan will be created and put into effect throughout the health centers. This intervention will also be incorporated into the standard auditing procedures. The anticipated duration for this phase is approximately 8 to 9 months, after which a second cross-sectional data collection will take place. The approach for this subsequent data collection will be consistent with that of the initial phase-*See Fig 1 for study flow chart*.

**Fig 1:**
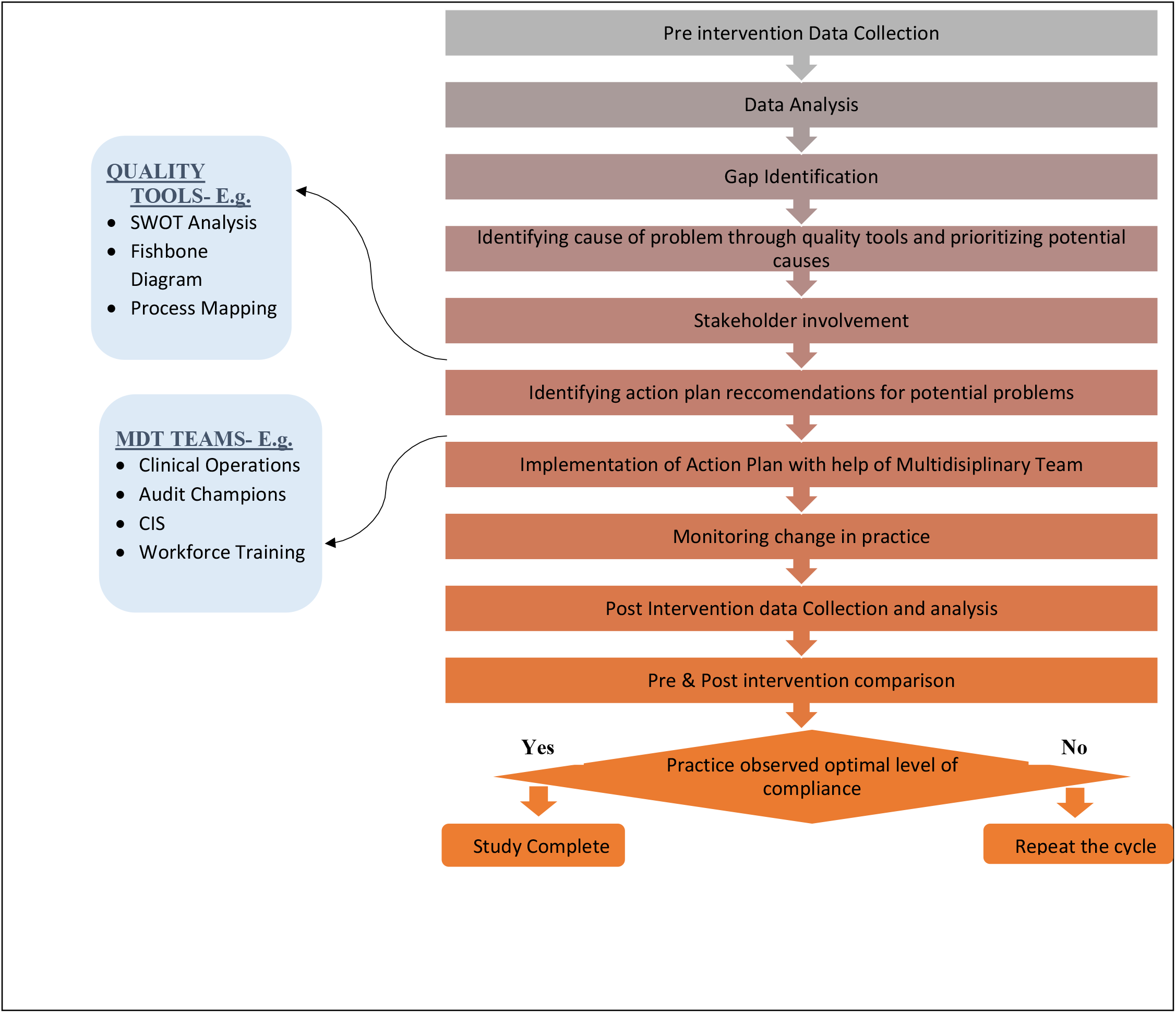
ICAE-DM CARE STUDY flow chart

### Analytical Plan

Data will be extracted from the electronic health record system at two separate time points: one time at a point before the intervention and the other time at a point after the implementation of the intervention. Excel will be used to collect and organize the data. PivotTables will be used to determine the frequency of cases across variables. Excel will be further used to produce histograms and other figures.

Study outcome variables will be assessed using independent tests like Chi square or t test to compare between the two groups. For satisfaction, we will do frequency by percentage P=(F/T) ∗100.

The statistical analysis will be performed using STATA 15.1 (College Station, TX, USA). A Wilcoxon sign rank test will be used to find the mean difference between the pre- and post-assessment. P<0.05 will be considered statistically significant. A logical model of protocol presented in *Fig 2*.

**Fig 2:**
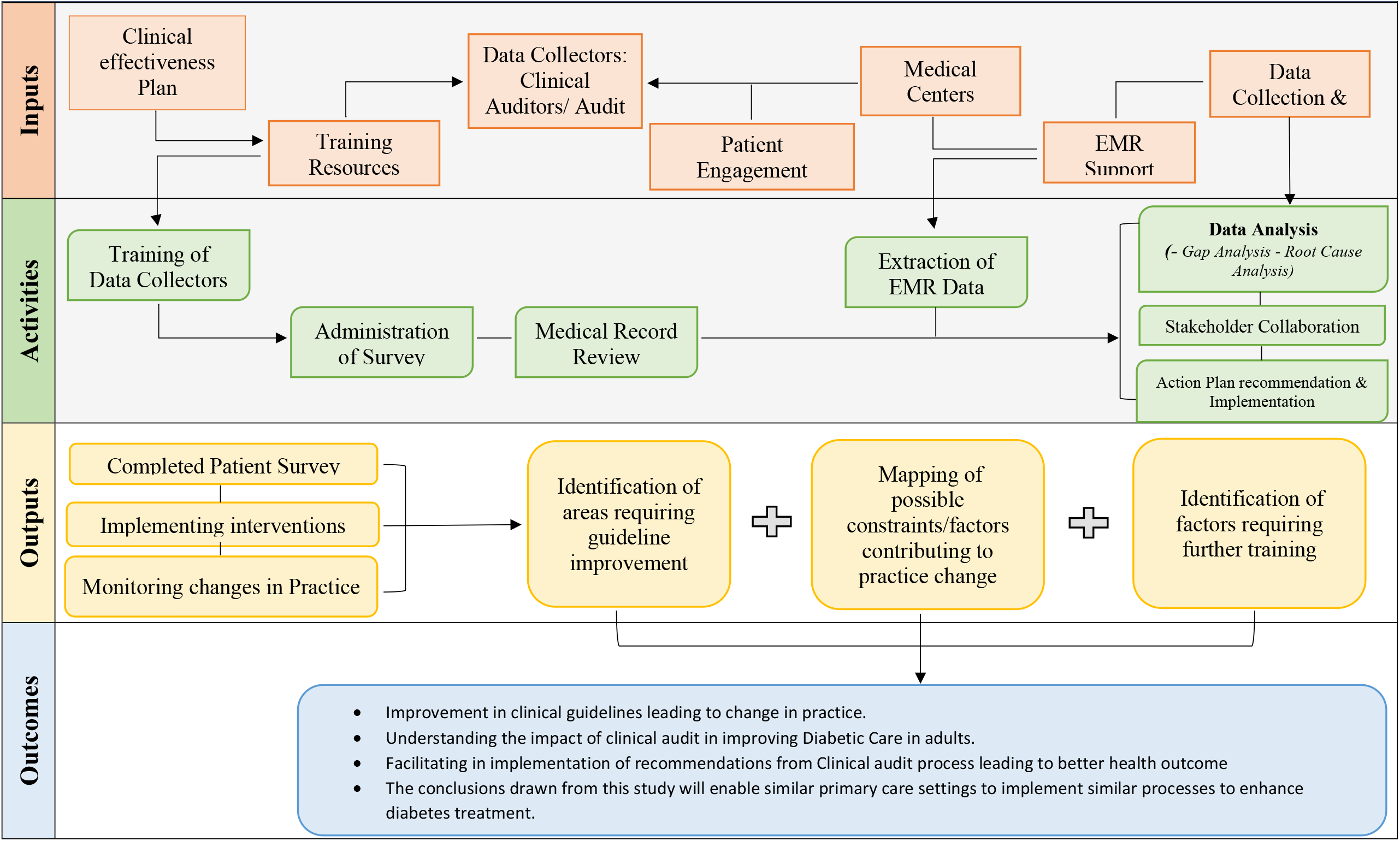
ICAE-DM CARE STUDY Logical Model

## Ethical approval

The Institutional Review Board (IRB) of PHCC (BUHOOTH-D-24-00059) has reviewed and granted approval for the study. Verbal consent will be obtained from participants who are 18 years of age or older for the telephonic survey. The overall planning and execution of the study will be conducted with integrity, in accordance with established ethical principles.

## Discussion

Continuous quality assurance efforts are fundamental for the assessment of clinical practices to ensure they are performed as intended. This is particularly important for conditions like diabetes, which require continuous management, whether in pre-diabetes or confirmed diabetes cases. Adherence to established guidelines is necessary to avert the progression from pre-diabetes to confirmed diabetes and to mitigate complications in individuals with confirmed diabetes. This study is designed to closely reflect real-world challenges in diabetes care at the primary care level and to evaluate how identified deficiencies have been addressed through targeted quality improvement initiatives. Additionally, the study will assess the impact of these interventions on clinical practice and determine whether the action plan led to significant improvements or any adverse outcomes. All scenarios will be analyzed in detail throughout the study.

The study will commence with a situational analysis of the current state of diabetes care, referred to as the pre-intervention phase. Data will be gathered using a specially designed data collection sheet, which will involve a review of medical records. Simultaneously, a telephonic survey will be conducted to gather insights regarding patients’ experiences with the diabetic care they have received. Following the data collection process, an initial analysis will be conducted. The analysis reveals deficiencies in current practices. The subsequent phase will involve identifying the underlying causes of the issues using various quality improvement methodologies, such as process mapping, fishbone diagrams, and focus group discussions. The choice of tools will be determined by the specific nature of the identified problem. This will be succeeded by interventions tailored to address the root causes identified. An effective action plan will be formulated utilizing SWOT analysis. The chosen interventions will be executed through a collaborative, multidisciplinary approach.

The multidisciplinary team is composed of the clinical effectiveness section, the clinical operations department, clinical audit champions at the health center level, and the clinical information system (CIS) to address any gaps identified concerning the electronic medical record system. This strategy will employ a three-tiered framework. To begin, a baseline assessment study report that outlines the identified gaps and intervention programs will be shared with the regional directors. The regional directors will then distribute this report to the health center managers, who will implement the action plan with the assistance of key stakeholders, including the physician lead, head nurse, pharmacy lead, and lab lead at the health center. The clinical audit champions will be responsible for monitoring and coordinating the implementation at the health center level.

The second-tier strategy entails the presentation of the action plan’s implementation progress at the taskforce meeting, where representatives from relevant departments gather. This meeting serves as a platform to address progress, identify challenges, and explore solutions, all with the shared objective of enacting change. The clinical effectiveness section will oversee the coordination of this meeting. Departments including clinical information systems (CIS), business health intelligence (BHI), workforce, and operations will be involved in the taskforce meeting. Each department will be accountable for executing the action plan within its area of responsibility.

The third-tier strategy involves a focal person from the clinical effectiveness section who collaborates with clinical audit champions at each health center within the region. These champions serve as an extension of the clinical effectiveness section at the health center level. They are authorized to implement changes with the backing of the health center manager. The advancement of the change initiatives will be reviewed during regular follow-up meetings, held every six weeks, among the focal points and champions. These meetings will be organized by region, during which champions will provide updates on progress, challenges, and other pertinent issues related to the implementation of changes at their respective health centers. The clinical effectiveness section will assist the champions in addressing any obstacles and will offer necessary guidance for the successful implementation of changes. This three-tiered approach will facilitate the implementation of changes. These activities will be carried out under the supervision of the clinical effectiveness section.

The post-intervention phase is scheduled to take place approximately 8 to 9 months following the initial study and the subsequent intervention. This timeframe will provide clinical staff with the opportunity to adapt to the new changes. The sample size for this study will mirror that of the first cross-sectional study. Data collection will be performed through an open chart review, similar to the initial study, with information being recorded on an Excel spreadsheet. We will gather the same data from the medical records as was collected during the first cross-sectional study. Upon completion of data collection, a final analysis will be conducted to evaluate the changes in the measured parameters. The comparison between pre- and post-intervention data will illustrate the impact of the clinical audit process on practice. The proposed protocol illustrates a solid quality improvement study that can be performed in a concise timeframe and with restricted resources yet features very effective and innovative interventions. This will be advantageous for healthcare systems looking to carry out similar studies in primary care settings and can be conducted more frequently to enhance the management of diabetes.

## Data Availability

All relevant data from this study will be made available upon study completion.

